# Bangladesh should engage the private sector for malaria elimination by 2030

**DOI:** 10.1101/2024.04.10.24305490

**Authors:** Ryan Patrick McArdle, Ching Swe Phru, Mohammad Sharif Hossain, Mohammad Shafiul Alam, Kasturi Haldar

## Abstract

Bangladesh reduced malaria by 93% from 2008-2020 through the action of governmental and non-governmental organizations, yet ∼18 million people continue to live at risk of infection. For-profit private healthcare providers, catalytic for malaria elimination in many countries, have not yet been integrated into the national program. By imposing strict definitions on a large and complex literature surrounding private healthcare sector engagement, we distilled eight distinct strategies important in other developing settings: contracting, financing, subsidization, regulation, social franchising, demand-side interventions, infrastructure-building and training. We weighed these in the context of Bangladesh’s flourishing private health care sector—driven by patient demand, self-interest, and aspirations for public good—as well as the heterogeneity in provider capacity and malaria prevalence across districts. We developed a new model dependent on five strategies of subsidization, training, infrastructure building, demand-side intervention, and referral financing, to empower Bangladesh’s phased agenda of eliminating indigenous malaria transmission by 2030.

## INTRODUCTION

Malaria remains one of the deadliest communicable diseases worldwide, claiming an estimated 627,000 lives in 2020 and leaving almost 50% of the world’s population at risk of contracting the disease. Despite this, malaria is treatable, and recent epidemiologic and pharmaceutical advances have allowed the complete elimination of malaria from 40 countries overall and 13 since the turn of the century.^1^

Among the remaining malaria-endemic countries, Bangladesh has experienced remarkable recent success in the control of malaria. Historically, Bangladesh suffered from a high malaria burden, and initial disease control efforts in the 1960s, which focused on the use of dichlorodiphenyltrichloroethane (DDT) to eliminate mosquito vectors,^2^ were disrupted by the cyclone of 1970, the Banglaeshi War of Liberation, and the international ban on DDT. In 2008, however, the National Malaria Elimination Program (NMEP) and a consortium of NGOs headed by the Bangladesh Rural Advancement Committee (BRAC) began revitalizing malaria control efforts within the country.^3^ This renewed strategy combined the mass distribution of long-lasting insecticide nets (LLINs) to reduce bites from malaria-transmitting mosquitos, in-home review of appropriate net use, rapid point-of-care diagnosis, and prompt treatment with frontline artemisinin-based combination treatments (ACTs; Fig. 1). This strategy has allowed the country to achieve near-elimination status in 8 of its 13 malaria-endemic regions (Fig. 2a and 2b);^3^ likewise, malaria case and death rates within the country decreased up to 2020 (Fig. 2c).^3^ This rapid progress prompted Bangladesh to set the ambitious target date of 2030 for the scaled elimination of malaria from the country.

**Figure 1.**
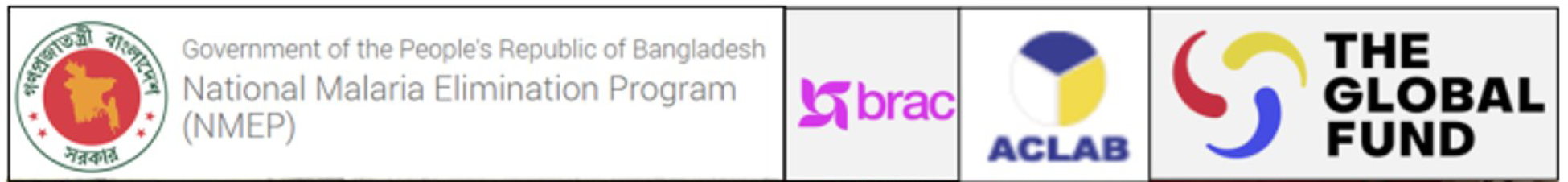
Malaria Elimination in Bangladesh. Supported by the Global Fund, the Govt of Bangladesh’s National Malaria Elimination Program (NMEP) works in conjunction with the NGO BRAC as well as others such as ACLAB (Alliance for Cooperation and Legal Aid Bangladesh). LLINs (long-lasting insecticide nets; indicated in (a) and (b)) are issued for use to prevent mosquito biting at night or rest periods. A paid, trained, high-school graduate, female health care community supervisor or Sasthya Kormi (SK; depicted seated in (b)) visits with a homesteader (Hstd; depicted standing in (b)) to ascertain appropriate use of nets and the number of family members protected under them and to remind Hstds of the longer-term maintenance of the LLIN over the expected 3-year life span. The SK also queries about recent fevers of and travel by family members. Reports of fever over the prior 24-48 hours are rapidly followed up by an in-home RDT. If negative, affected individuals are given a written referral to a local health clinic for further follow up. Severe patients are immediately sent to a suitable clinic or hospital. Patients positive for *P. falciparum or P. vivax* with uncomplicated malaria are directly started on appropriate antimalarial drug treatment by the SK. The SK keeps contact via cell phone at the times of all subsequent doses (∼8 h apart) to check on the patient and compliance. In addition, she visits in-person each day that the patient continues to be febrile. Conversely, febrile or otherwise-ill patients also directly call their SK for an immediate home visit, RDT, drug treatment and collection of clinical history and travel data. All this information as well as the spent RDTs are transferred to the NGO and NMEP. 80% of all malaria infections in Bangladesh are identified by SK’s or Sasthya Sebikas (who report to SK’s and do not administer antimalarial drugs unsupervised). The result is high-quality, accountable, patient-centered care, with collation of rich natural history data at a national scale to inform the malaria elimination agenda, as well as empowerment of community health workers who are aware of the central role they play in Bangladesh’s success in diminishing malaria as well as supporting the general health of their communities.

**Figure 2.**
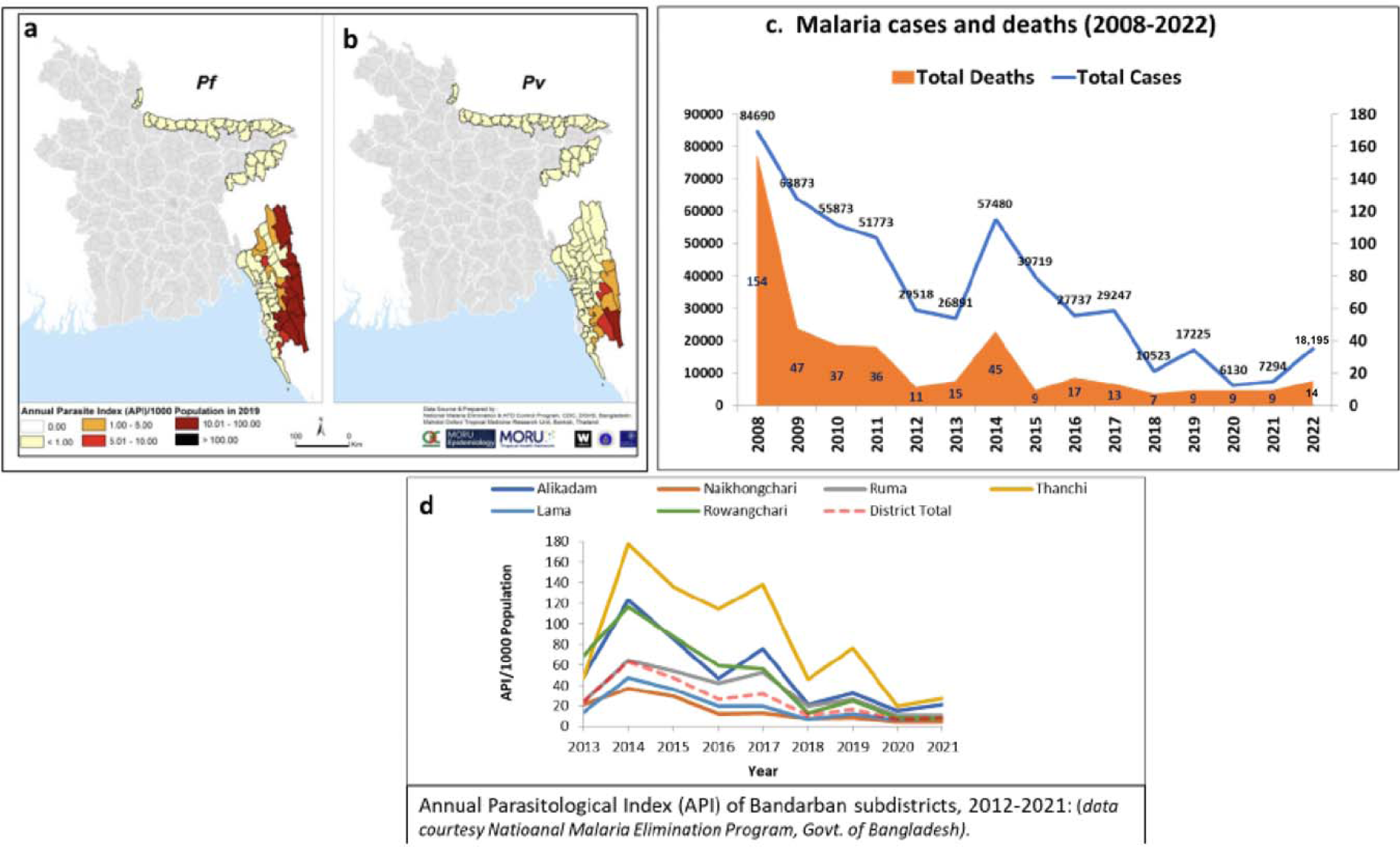
Dynamics of malaria control in Bangladesh. a-b, Phased malaria elimination in Bangladesh 2010-2030 for P. falciparum (Pf) and P. vivax (Pv); c, Malaria cases and deaths in Bangladesh 2008;-2020 d, Annual Parasitological Index (API) 2013-2021 in sub-districts of Bandarban, the CHT district with highest levels of malaria. (All data are courtesy of the National Malaria Elimination Program, Govt of Bangladesh).

However, several challenges persist. Three high-transmission districts (Bandarban, Rangamati, and Khagrachari) remain in the Chittagong Hill Tracts (CHTs; Fig. 2), a region which in 2021 and 2022 showed increases in malaria incidence (Fig. 2c-d), posing significant challenges for continued elimination efforts.^3^ These districts are poorly served by roads and transportation, which are necessary for the national program’s healthcare workers to comprehensively access homes, deliver point-of-care diagnosis and treatment, and ensure proper use of LLINs throughout all subdistricts. These districts also feature a high proportion of remote communities of isolated, Indigenous (non-Bengali) peoples, presenting cultural practices that compound geographical barriers to prevent access for malaria elimination programs.^4,5^ In contrast, two low-endemic districts, Cox’s Bazar and Chittagong, are more accessible to present control efforts. These regions plan to eliminate the endogenous transmission within the districts by 2025, but will continue to face the prospect of imported malaria due to high levels of human travel from the neighboring high-transmission CHT districts until the 2030 elimination date.^6^ Eight northeastern districts at near-elimination present only a few cases of malaria annually. Here, there is concern that because risk of infection is lowered, LLINs are not consistently utilized. LLINs also may not protect agricultural workers in tea gardens.^7^

One important task for the malaria elimination program in Bangladesh is to better involve the forprofit private healthcare sector into malaria elimination efforts. So-called private sector engagement (PSE) has in recent years become a point of emphasis within both regional and global malaria elimination programs due to the large proportion of patients who seek care from private healthcare providers (PHPs).^8,9^ This high utilization of the private healthcare sector holds true also in Bangladesh, where in some endemic regions private healthcare providers represent over 97% of all healthcare available.^10^ Global trends in the epidemiology of malaria and other diseases near or at elimination, such as draculaniasis and smallpox, suggest that as malaria elimination in Bangladesh advances, an increasing fraction of disease cases will begin to localize amongst difficult to reach subpopulations; as PHPs oftentimes have greater reach than public entities, it will be essential to find ways to include them in the broader movement towards malaria elimination within the country.^11–13^ In addition, current data from Bangladesh finds that only 80% of malaria diagnosis and treatment in Bangladesh occurs within the national program (see Fig. 1), suggesting as much as 20% of patients seek care from PHPs. While malaria is a notifiable disease in Bangladesh,^14^ many PHPs are unaware of this fact, resulting in serious underreporting; this poses a barrier to WHO certification of malaria elimination, which requires proof of a comprehensive surveillance system.^15^ In addition, untrained private providers, who often prove difficult to regulate, have been shown to provide ineffective management of malaria cases in other developing settings,^16^ threatening to increase both malaria deaths and antimalarial resistance. Thus, in order to meet the 2030 target date for malaria elimination, malaria control efforts in Bangladesh will require robust incorporation of PHPs into existing surveillance networks, granting PHPs the ability to aid in case detection and requiring them to refer confirmed-positive cases to public treatment frameworks.

This policy review aims to collate and analyze the strategies used for PSE elsewhere in the literature, weighing them in the context of challenges facing malaria elimination in the high-transmission districts of the CHTs as well as in the low-transmission and near-elimination districts, and develop a multi-modal plan to suggest the most promising path for PSE in Bangladesh.

## METHODS

### LITERATURE REVIEW

To the extent that the literature allowed, this study limited its scope to low- and middle-income countries (LMICs). The study also focused on efforts to engage the for-profit private sector in healthcare delivery at the local level; studies on the engagement of larger international private entities, such as drug manufacturers, were largely excluded. Primary concern was given to literature on PSE directly in the context of malaria elimination and to studies in a Bangladeshi context; however, due to a relative absence of studies for many strategies, evidence from other health initiatives and settings was consulted, including grey literature. This study also excludes data on the BRAC-led NGO consortium; as indicated earlier, BRAC and its subsidiary NGOs have been closely integrated into modern public malaria elimination efforts from their inception, and their status as nonprofit entities causes them to respond to different factors than for-profit PHPs. Due to the breadth of literature on private sector engagement as a whole, this paper also gave preference to existing literature reviews where possible over a review of the primary literature itself.

Sources were discovered using the search tool of Google Scholar; Google Scholar was selected over other databases due to its ability to probe both white and grey literature. Preliminary research revealed several distinct strategies for private sector engagement: “contracting”, “financing” (including “pay for performance”, “rewards system”, and “referral system”), “subsidization”, “franchising” (including “accreditation” and “certification”), “regulation”, “demand-side” interventions (including “social marketing”, “public awareness campaigns”, “mass media campaigns”, and “community mobilization”), “infrastructure-building strategies” (including “public-private partnership”), and “training”. Each of these areas was thoroughly queried; full search terms are available in the supplemental materials. Only source published in English were reviewed. Following the guidelines listed in Bramer 2019, 100 sources were included for each collection of search terms before deduplication.^17^ Identified sources were also supplemented by lists of sources derived from existing review papers.^9,18,19^

### THE BANGLADESH CONTEXT

The Bangladeshi context was initiated through a combination of published literature in Bangladesh, insights from informal conversations with malaria elimination experts from NMEP and BRAC, and focus group discussions with private healthcare Providers.^3–7,20^

## RESULTS

### STRATEGIES FOR PRIVATE SECTOR ENGAGEMENT BASED ON MOTIVATION

As summarized in Fig. 3, literature searches performed between October 1, 2022 and January 31, 2023 revealed 3,026 sources. Of these results, 818 were duplicates, leaving 2,208 sources after deduplication. Of these, 235 sources (including full-length articles, NGO and governmental reports, and books) were reviewed and included in this policy review (a full list of all sources can be found in the supplemental materials). Since the high transmission CHT districts have the highest need for new approaches to reduce malaria, we queried the data to find that only seven of 235 sources referenced CHTs and even those did not provide in-depth accounts of strategies used to engage PHPs; this suggests that an assessment of PSE strategies reported elsewhere and an application of lessons learned to the engagement private malaria providers in the CHTs is an area of high unmet need. Indeed, our literature search revealed zero sources analyzing the engagement of for-profit private sector healthcare providers with the NMEP in the CHTs (Fig. 3). Similarly, we were unable to find studies reporting on PSE in low-endemic and near-elimination districts. We therefore assessed the remaining 235 sources, identify eight distinct strategies of contracting, financing, subsidization, social franchising, regulation, demand-side interventions, infrastructure-building strategies, and training (Fig. 4). A review of the Bangladeshi private healthcare sector revealed three primary motivators for private healthcare providers: provider self-interest, patient demand, and altruism (concern for patient good). Four mechanisms (reward, punish, building infrastructure, building knowledge) that connect strategies to motivation are also shown.

**Figure 3.**
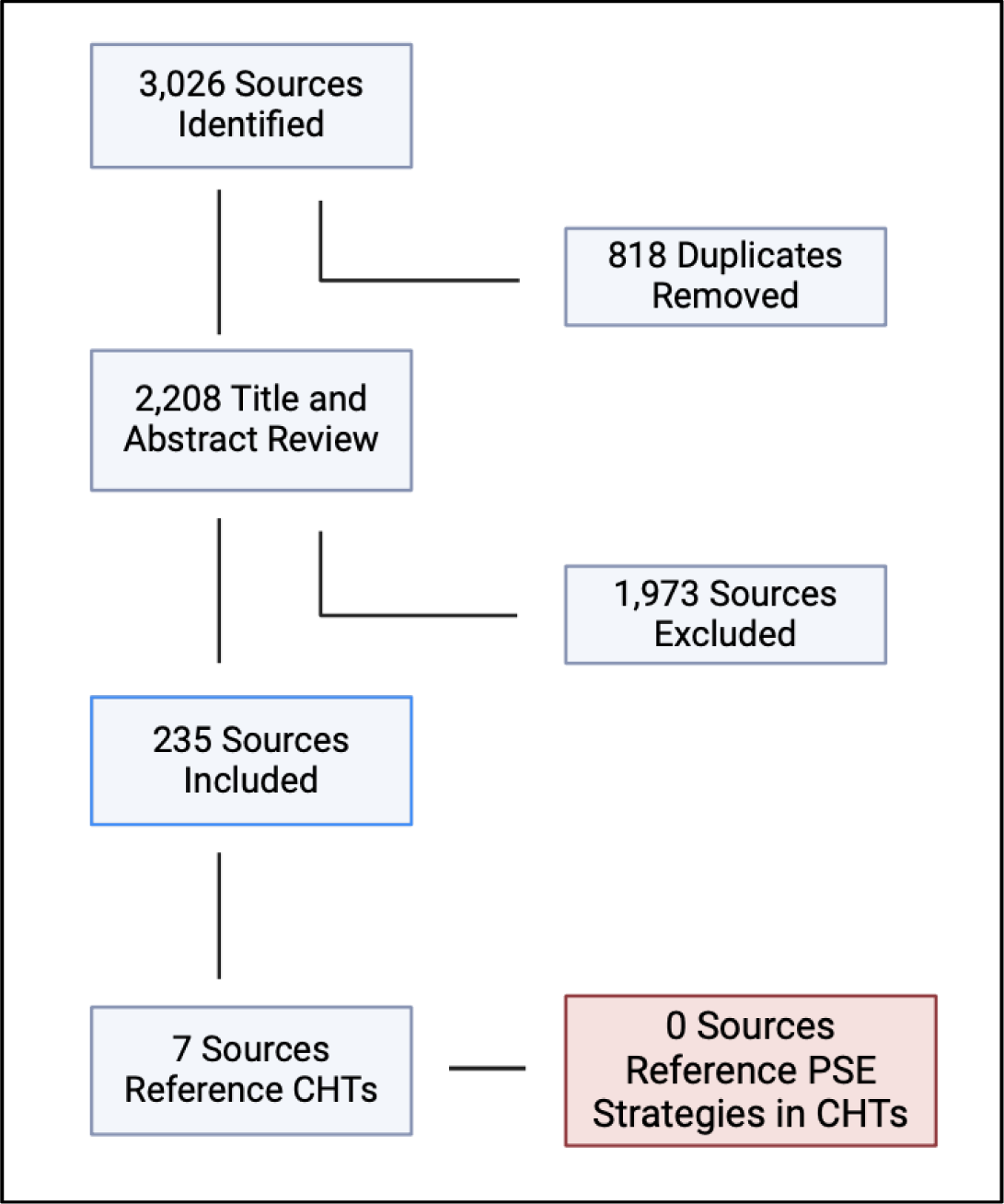
Absence of Literature on PSE in the CHTs. A literature search was conducted with Google Scholar to analyze the state of literature on private sector engagement (PSE) in the Chittagong Hill Tracts (CHTs). Searches of the white and grey literature revealed 3,026 sources, from which 818 duplicates were removed, leaving 2,208 deduplicated sources which proceeded to title and abstract screening. Of these, 1,973 sources did not meet the inclusion criteria and were excluded, leaving 235 sources which were included in the study. Further analysis revealed that only 7 of these sources referenced the CHTs, and none of those referenced for-profit PSE strategies in the context of the CHTs, demonstrating an area of high unmet need in the literature. Created using biorender.com.

**Figure 4.**
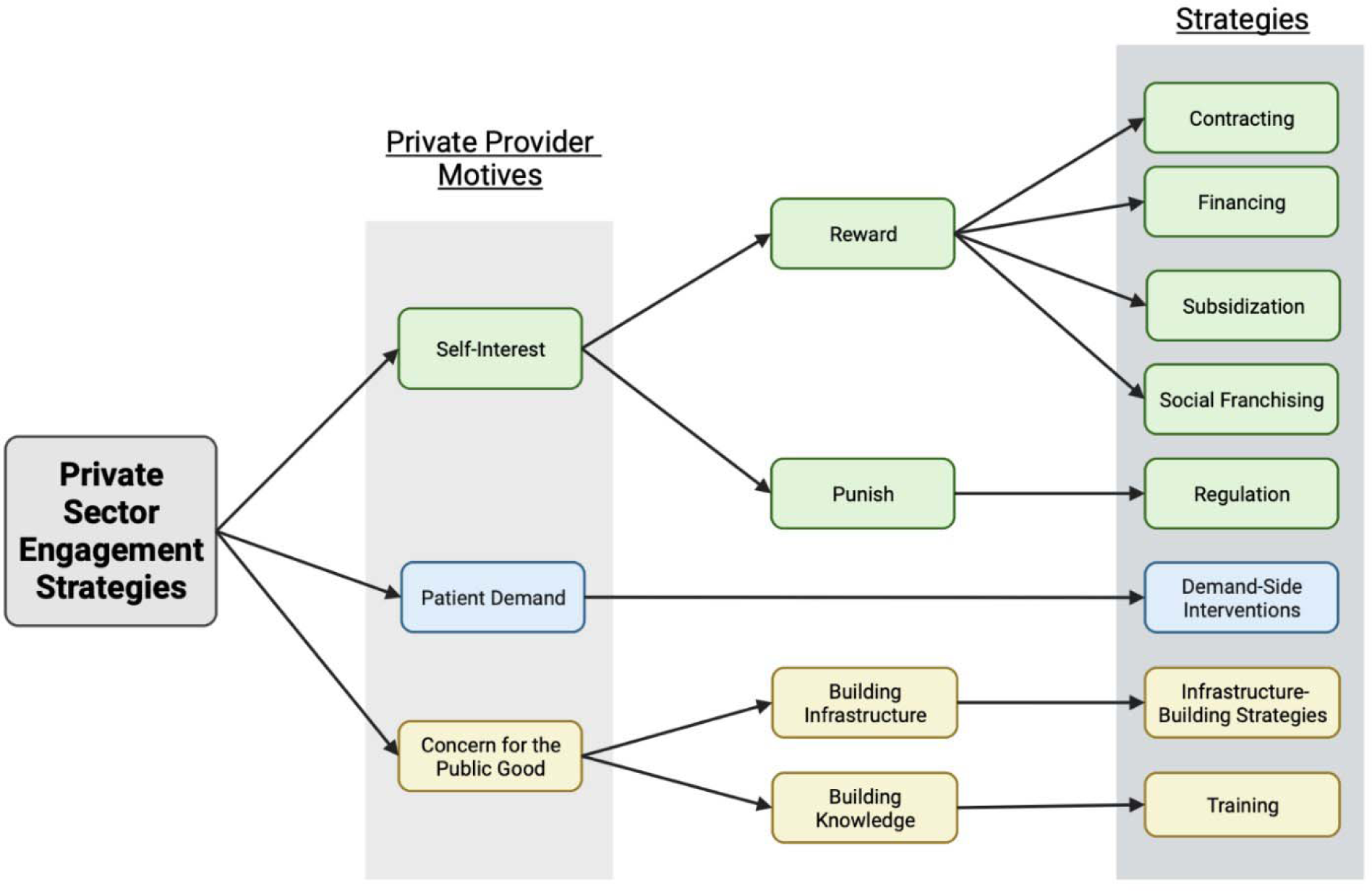
Potential PSE Strategies for PHPs in Bangladesh. Based on a review of private sector engagement (PSE) in the literature (see Methods and Fig. 3), 8 potential strategies for PSE in the context of malaria elimination were revealed: contracting, financing, subsidization, social franchising, regulation, demand-side interventions, infrastructure-building strategies, and training. A review of the Bangladeshi private healthcare sector revealed three primary motivators for private healthcare providers: provider self-interest, patient demand, and altruism (concern for patient good). Four mechanisms (reward, punish, building infrastructure, building knowledge) that connect strategies to motivation are also shown. Created with biorender.com.

In Fig 4, the first set of strategies appeal to the self-interest of private providers. Assuming that PHPs lack adequate incentives to comply with public health mandates, these strategies seek to impose either punishments (“regulation”) or rewards (“contracting”, “financing”, “subsidization”, and “social franchising”) to alter the behavior of PHPs driven by self-interest. The second set of strategies target the well-documented reliance of PHPs on patient demand. By seeking to modify patient demand for specific services, these so-called “demand-side interventions” act as indirect modifiers of provider behavior. The final set of strategies address inherent concern for the public good among PHPs. Unlike the other strategies, this category assumes that providers have inherent motivation to aid in public health programs but lack the ability to do so. By building either technical and organizational infrastructure (“infrastructure-building strategies”) or knowledge (“training”), these strategies aim to empower previously-motivated PHPs to aid in disease elimination efforts.

### HETEROGENEITY IN THE BANGLADESH PRIVATE HEALTHCARE SECTOR

Though relatively unstudied, evidence suggests that Bangladesh features a large private healthcare sector. The Bangladeshi private healthcare sector is also heterogenous, marked by the presence of both formal and informal providers. Formal providers, who have been trained and registered through official government channels, include private hospitals and private healthcare provided to employees by corporations; on the other hand, informal providers, who either have no formal medical training or have not registered their practice with the government, include untrained drug vendors, traditional healers such as religious, ayurvedic, and homeopathic therapists, and even formally-trained physicians who have unregistered clinics in addition to their public practice.^9,21^

Several recent qualitative studies on the motivations of PHPs in Bangladesh and in Southeast Asia more broadly provide excellent insights into the environment in which the Bangladeshi private healthcare sector operates.^20,22^ PHPs in Bangladesh must compete with cheaper but often overcrowded public healthcare options; as a result, the for-profit private sector finds a competitive advantage in its high responsiveness to patient demand and prioritizes the maintenance of strong personal relationships with patients. The for-profit private sector in Bangladesh also features a rich referral network, both from the public sector and between providers in the for-profit private sector; referral fees, which constitute a financial compensation paid to the referring practitioner, are common practice. Pharmaceutical representatives are also reported to have an outsized influence on the products available in local drug stores and on the prescription practices of private physicians due to their widespread provision of incentives, including direct payments and the provision of free product, to PHPs. In summary, PHPs in Bangladesh operate within a highly informal, complex marketplace, and their decisions are driven by a combination of internal and external factors including self-interest, patient demand, and, in some cases, concern for the public good.^20^

### ASSESSMENT OF EIGHT STRATEGIES OF PSE IN THE BANGLADESH CONTEXT

#### CONTRACTING

Contracting entails the direct employment of private healthcare providers (PHPs) for the delivery of specific healthcare services. In contracting, PHPs are usually hired as full-time employees who exclusively perform contracted activities.^23^ Despite strong theoretical appeal driven by the perceived efficacy of the market, contracting for healthcare delivery has in practice had mixed results. Although several individual studies and one review reported positive outcomes,^23–25^ multiple systematic reviews have found that existing positive evidence for contracting is low-quality and that when proper study design was followed, there was little evidence that contracting programs have any effect on targeted health outcomes.^19,26–28^ Nevertheless, even critics acquiesce that contracting remains a feasible strategy when used to expand health services in settings where existing public health infrastructure is unable to meet patient demand.^27^

Contracting represents a poor solution in the Bangladeshi context. Bangladesh already features a comprehensive public system for malaria treatment, negating the positive effects that contracting can have when used to expand healthcare access. In addition, malaria cases in Bangladesh are too low to justify hiring networks of private providers as full-time malaria workers, leaving contracting strategies financially infeasible.

#### FINANCING

Like contracting, financing provides direct financial incentives as a mechanism to spur private sector engagement; however, instead of fully employing PHPs for a salary, financing efforts simply incentivize desired outcomes by providing smaller monetary rewards to PHPs.^29^ Financing, which is synonymous with “pay-for-performance” (P4P), has been used widely and features an extensive body of literature. Despite this, evidence for the success of P4P programs as a whole is mixed, both amongst individual studies and reviews.^19,30–36^ However, evidence does seem to suggest certain criteria shared by successful P4P programs. In general, highly tailored P4P programs which use a carefully-measured incentive to target specific providers and healthcare outcomes have enjoyed higher degrees of success;^31,34,35^ successful programs have also taken particular care to integrate routine evaluation in order to avoid the potential unintended consequences of incentivization.^31,33,34^

In the context of disease elimination, P4P strategies can be used to incentivize the reporting of disease cases to centralized surveillance networks. Practical applications of such rewards systems suggest that they can be effective, albeit often through the use of low-quality evidence.^12,37–40^ In one of the stronger pieces of evidence, a systematic review of several PSE strategies reported that referral rewards systems represented the most effective strategy studied and resulted in positive changes in provider behavior 84% of the time.^18^ Rewards systems must be accompanied by appropriate strengthening of infrastructure, as timely repayment is vital to PHP buy-in;^41^ nevertheless, they represent a potentially useful solution for disease programs.

Financing has high potential as a mechanism for incentivizing small, informal providers in Bangladesh. In particular, referral fees, or the provision of a small financial reward for the referral of malaria-positive patients whose diagnosis is confirmed at public clinics, represents a highly targeted, direct manner of incentivization. Such a system could be structured similarly to that reported in Vo et al., where PHPs in Vietnam were given a cash reward for reporting confirmed-positive tuberculosis cases to the public surveillance system.^40^ In fact, the provision of referral fees is already standard practice amongst members of the Bangladeshi private healthcare sector, as it helps the referring PHP to offset the lost revenue from treating a patient. Because of this, such a referral reward system may be necessary to achieve any desirable degree of compliance with government referral mandates.

#### REGULATION

Rather than providing incentives for compliance with public health objectives, regulation functions by imposing legal punishments for noncompliance.^21^ The success of regulation largely depends on the capacity for enforcement.^42^ Where enforcement networks are strong, regulation has generally proven successful;^37,43^ more often, however, regulatory strategies in developing settings face much weaker enforcement support and, as a result, much more modest results.^42–46^ While limited, evidence suggests that regulation of PHPs has been similarly ineffective in Bangladesh.^47^ Regulatory strategies face the additional risk of unintended consequences; in particular, highly stringent regulations threaten to reduce healthcare access for vulnerable populations.^45,48^

Other regulatory efforts have focused on “self-regulation”, an arrangement in which the powers and duties of enforcement are delegated to regulatory bodies composed of members of the for-profit private sector.^46^ Despite appeal as a hands-free mechanism for enforcement,^45^ self-regulatory efforts have been equally ineffective, largely because of both a lack of proper resources and “regulatory capture”, a well-documented phenomenon in which self-regulatory boards use their power for personal benefit.^19,44–46^

Although malaria is a public health notifiable disease in Bangladesh,^14^ regulation is unlikely to form a successful strategy for PSE within the country; in addition to inefficacy in other developing settings, current regulations in Bangladesh are complicated by poor enforcement networks, an issue which would only be further complicated by the geographical remoteness of remaining malaria hotspots.

#### SUBSIDIZATION

Subsidization provides free or discounted medications or supplies to PHPs to overcome the cost barriers that may prevent their full engagement with public health priorities. Subsidization provides no direct positive incentive to PHPs, instead removing cost barriers that can present negative incentives to engagement. Subsidization has been used widely in the context of malaria control and elimination efforts, both for direct treatments (i.e. artemisinin combination therapies, or ACTs) and for diagnostic methods (i.e. rapid diagnostic tests, or RDTs). Most evidence for ACT subsidization has come from the Affordable Medicines Facility-malaria (AMFm) program, an international cooperation which allowed pharmaceutical wholesalers in developing countries to purchase ACTs directly from manufacturers at reduced cost.^49^ Despite concerns of negative residual effects from some commentators,^50^ the AMFm program has proven largely successful at increasing access to and use of ACTs.^9,49,51^

RDT subsidization efforts have been fewer in number, and their results have been more mixed. Both individual studies and a systematic review provide unclear evidence as to whether RDT subsidies represent an effective mechanism for either reducing antimalarial over-prescription or increasing RDT access and usage, although results for both were generally better when RDT subsidies were higher.^52–55^ However, other commentators have argued against large subsidies, noting that the subsidized RDT price must preserve the ability of private providers to generate a reasonable profit.^56^

Overall, this data suggests that subsidization efforts are perhaps best evaluated on a case-by-case basis and must take particular caution to fully investigate both the incentives facing providers and the potential unintended consequences of interventions. Nevertheless, subsidization, and in particular the subsidization of RDTs, presents a promising solution for Bangladesh. Subsidized RDTs could help to encourage PHPs to test a greater proportion of their patients, and subsidizing high-quality, WHO-certified tests could help to crowd out lower-quality RDTs from the market.

#### SOCIAL FRANCHISING

In social franchising strategies, for-profit private sector providers bear the brand of the franchising organization, agreeing to operate under some sort of regulatory structure and in some cases to pay fees to the coordinating body. In return, franchises offer PHPs both the “hard” incentive of access to cheaper, higher-quality drugs and the “soft” incentives of increased credibility from accreditation or certification programs and increased clientele volume through marketing strategies.^57^

Evidence from a variety of health initiatives suggests that social franchising efforts have been successful in improving the quality of care of private providers, client satisfaction, and client knowledge of health conditions.^19,58,59^ However, social franchising strategies as a whole have struggled to improve health outcomes and healthcare utilization on a community level due to their increased price.^45,58–60^ This shortcoming makes social franchising strategies as a whole particularly poorly suited for malaria elimination in Bangladesh, where the majority of cases are concentrated amongst rural laborers.

However, one component of social franchising strategies may be particularly relevant to malaria programs in Bangladesh: certification. In certification strategies, an organizing body grants PHPs some sort of endorsement for meeting specific guidelines or standards.^19,61^ Evidence for independent accreditation and certification programs is generally positive, and seems to support the assertion that accreditation and certification can improve quality of care.^19,46,62^

In the Bangladeshi context, certification presents a promising incentive for PHPs to receive training and comply with guidelines. Informal private providers are oftentimes eager to gain credibility within the community,^46^ and the increased authority from an official NMEP certification could present a valuable incentive for PHPs to attend training sessions and comply with public health guidelines.

#### DEMAND-SIDE INTERVENTIONS

Perhaps the largest determinant of PHP behavior is consumer demand; in order to maintain clientele in spite of cheaper public healthcare options, PHPs must carefully align their services with patient desires. As a result, demand-side interventions, which seek to modify the healthcare-seeking behavior of patients, represent a promising solution to change the incentives, and thus the behavior, of PHPs.^18,20,45^ Demand-side interventions take a number of forms, including social marketing, public awareness campaigns, mass media campaigns, and community mobilization. Because these categories present a fair degree of overlap and a comprehensive review of each is beyond the scope of this paper, their efficacy will be analyzed collectively.

Most evidence for demand-side interventions is weak; despite this, when taken together, evidence seems to support the potential for demand-side interventions to improve patient knowledge and awareness and to modify patient behavior, potentially improving demand for health services.^19,34,63–68^ While standalone demand-side interventions have struggled to improve health outcomes, most patient-facing strategies do nothing to improve healthcare quality.^63,64,68^ Characteristics associated with successful demand-side interventions include sufficient exposure to patient communities,^66^ careful integration with the existing context,^67^ and continual reinforcement.^68^

Demand-side interventions have been less-studied in the context of private sector engagement. While it is generally well-established that demand-side interventions can improve patient demand for specific health services and that private providers are highly responsive to patient demand, only a small, albeit positive, body of research supports the direct effects of demand-side interventions on PHP behavior.^18,30,65^ Nevertheless, demand-side interventions pose a promising strategy for malaria elimination efforts in Bangladesh, particularly when combined with certification programs: if demand-side interventions could successfully convince patient communities to value the certification, it would only further improve the likelihood that private providers would value certification. Demand-side interventions would have the added benefit of raising the public profile of malaria elimination efforts and helping to ensure patients would follow through with referrals to public clinics.

#### INFRASTRUCTURE-BUILDING STRATEGIES

For providers motivated by a concern for the public good, PSE may be as simple as building greater avenues for communication and cooperation between public and private entities; this paper will refer to these efforts as “infrastructure-building strategies”. Some of the most common manifestations of infrastructure-building are public-private partnerships (PPP). In the context of disease elimination, PPPs normally entail agreements between public health institutions and large businesses in which both parties agree to share the costs and burdens of disease elimination efforts, including vector control, testing, and treatment.^69^ Businesses have inherent motivation to engage in public health programs, benefiting both directly from a healthier workforce and indirectly from the increased “social license” that arises from helping to tackle health issues in local communities;^69^ as a result, businesses are often willing to share the costs of disease elimination efforts. Despite a high risk of bias due to funding sources, most PPP programs report sweeping successes.^69,70^

Beyond its use as a primary form of engagement with large businesses, infrastructure-building can be thought of more broadly as a foundational strategy for all other PSE efforts on an as-needed basis. Successful PSE programs require both technical support, such as supply chain integration and the construction of novel data platforms for case reporting, and organizational support, such as forging strong connections with local leadership.^31,67,71^

In the Bangladeshi context, infrastructure-building strategies present an appealing and necessary intervention. Where large industry is present, such as in the low-transmission and near-elimination districts, companies could be asked to either provide testing and refer cases to the National Malaria Elimination Program themselves or house an NMEP-trained health worker. In addition, given the variation in social and economic conditions across malaria-endemic districts, special consideration should be given to ensure that all PSE efforts in Bangladesh are accompanied by adequate technical and organizational support.

#### TRAINING

Even in situations where PHPs have the motivation and required infrastructure to support public health programs, they oftentimes lack the skills and knowledge needed to successfully comply with public health mandates;^72^ this is especially true in Bangladesh, where many PHPs operate informally and lack formal medical education. Training seeks to fill this gap, providing PHPs with the technical tools needed to aid in disease control efforts. Because of the clear necessity of an educated workforce, training is the most popular of all PSE strategies, representing over 70% of all interventions by some estimations.^9,18^

Evidence for the impact of independent training programs has been largely mixed.^18,29,72–75^ There is some evidence that pre-service training, which occurs before a PHP begins rendering service, is more effective than in-service training;^72,73^ as a whole, however, there is little agreement on whether training initiatives on their own can have a significant impact on PHP behavior. One potential explanation for this heterogeneity is the common admission that training initiatives work best in combination with other strategies and are best viewed as a complementary measure and can lead to positive outcomes when properly implemented.^9,18,29,45,72,73^ Key elements of successful training initiatives include continual reinforcement,^72,73^ well-trained trainers,^18,73^ and interactive training programs.^73^

Training represents an essential complementary strategy to any PSE efforts in Bangladesh. The informal nature of many private providers and high rate of turnover amongst PHPs necessitates frequent training and informational campaigns targeted towards providers. If PHPs are asked to test patients and refer cases, training on both proper testing methodology and case referral practices will be necessary to ensure the success of those programs. Training could also be easily accompanied by the provision of a certification to increase its apparent value to PHPs.

### RECOMMENDED MODEL FOR PRIVATE SECTOR ENGAGEMENT IN

#### BANGLADESH

Based on our findings, we recommend a two-step, multimodal model to optimize for-profit private sector engagement in Bangladesh (Fig. 5). As phase one of our intervention, we recommend the provision of free or subsidized RDTs to PHPs to ensure adequate testing and the training of providers on both proper RDT usage and case reporting (preferably through electronic reporting methods); successful completion of both by PHPs may be accompanied by the provision of an official certification. These initial steps, which should be simpler to implement, address the urgent nationwide need for greater knowledge and usage of current testing and referral protocols. As a second phase, and pursuant to the outcomes of the first, we recommend broadening campaigns to increase patient demand for providers with a government-sponsored certification, the provision of cash rewards to PHPs upon the referral of a confirmed positive case, and a continuing strengthening of the partnerships and logistics required to ensure program success.

**Figure 5.**
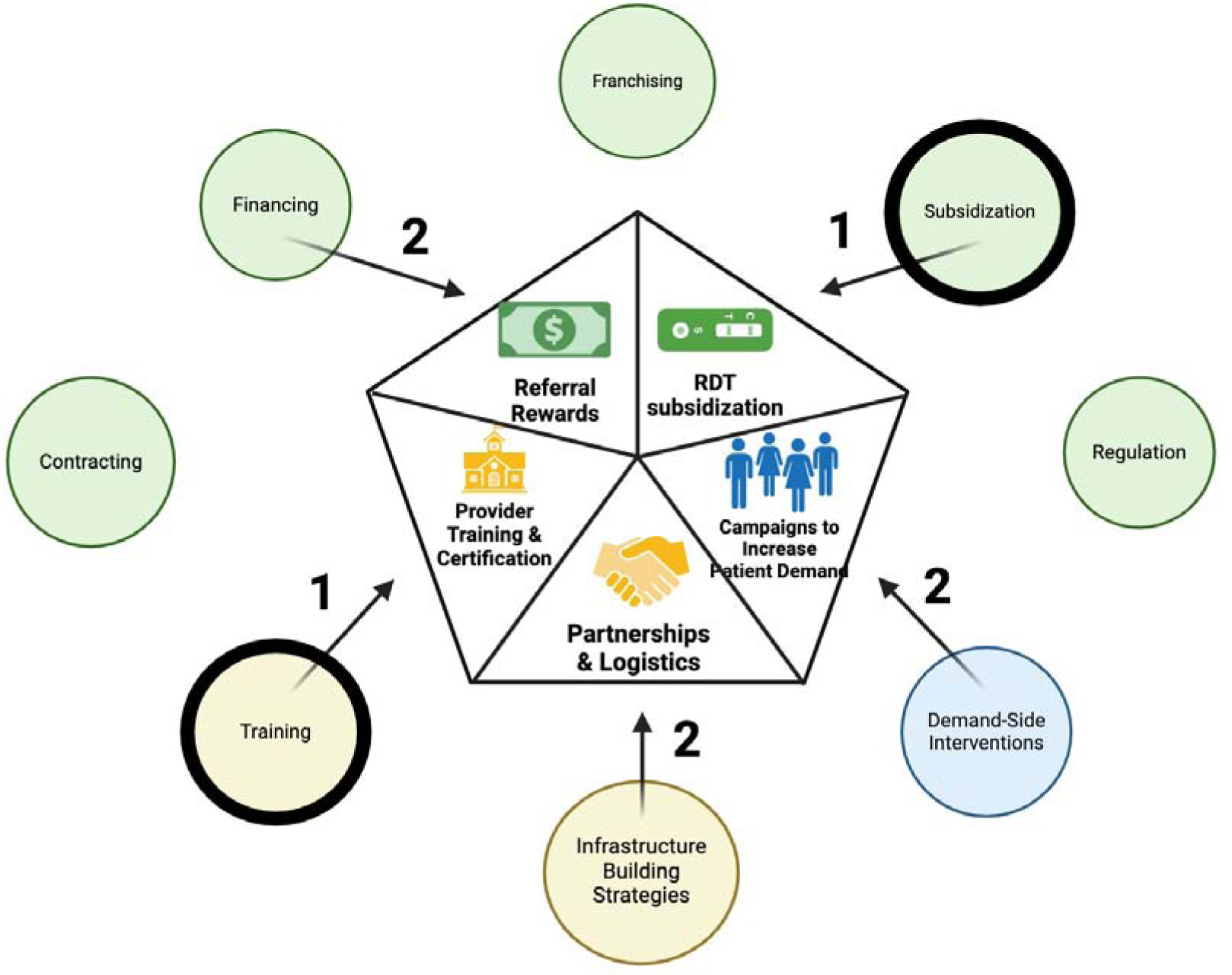
Model for PSE in Bangladesh. Matching the analysis of the success levels of individual strategies in the literature with the context of malaria elimination in Bangladesh, we created a stepped, multi-modal recommendation model for Bangladesh. This model targeted all three modes of provider motivation, including referral rewards financing and RDT subsidization for providers motivated by self-interest; campaigns to increase patient demand for providers motivated by patient demand; and training and the formation of partnerships and logistics for providers motivated by concern for the public good. The stepped feature accommodates phased elimination strategy of the National Malaria Elimination Program and conditions in high endemic districts of the CHTs, where there is urgent need to decrease malaria levels but the number of private providers maybe lower and a larger majority may be informal providers. Here close attention should be paid to the first step (1): training (of providers to conduct diagnosis with) subsidized RDT (both strategies are encircled in black bands) as well as enter data into formats needed by the national program. Success in utilization of RDT could be followed (2) with infrastructure support that can be supported by local resources of providers or the national program as well as referral financing and demand side intervention, to support networks that emerge between the patient community, private provider and NMEP. Created with biorender.com.

We recommend this stepwise but multimodal model due to the strength of evidence supporting multifaceted PSE programs.^8,9,29,45,66,75^ In one systematic review of strategies oriented towards informal PHPs, Shah et al. 2011 note that the average percentage of positive outcome measures rises steadily as the number of strategies employed by a program increases, doubling from 44% for interventions that used only a singular strategy to 88% for interventions that use 4.^18^ A multimodal model is also necessitated by the heterogeneity of motivation amongst PHPs in Bangladesh, all of which are targeted under our model.

While our findings suggest this general model for Bangladesh, the subdistrict and district-wise context will likely necessitate a phased application in the different malaria-endemic regions of the country. In the high-endemic districts of the CHTs, where there is urgent need to decrease malaria levels but the number of private providers maybe lower and a larger majority may be informal providers, immediate focus should rest on training providers to conduct diagnosis with RDTs and enter data into formats needed by the national program. Successful utilization of RDTs should be followed by infrastructure-building, and in particular efforts to improve connections with local leadership; stronger relationships between local leaders, PHPs, and the NMEP will be required to successfully target patient demand. Additionally, certification of trainers after proven performance could serve as a pre-requisite for the receipt of referral financing. In sum, malaria elimination efforts in the CHTs should focus on efforts to build both technical and organizational infrastructure in the immediate term.

In low-endemic regions such as Chakaria and Cox’s Bazar and the near-elimination settings of the northeastern districts, infrastructure is already sufficient to ensure that the other components of the model will have the logistical support and local partnerships necessary for success; as a result, the application of the other portions of the model in these regions should be able to proceed relatively unimpeded.

#### SUMMARY

For-profit private sector healthcare providers have not been incorporated into Bangladesh’s public-sector malaria elimination efforts, namely the National Malaria Elimination Program and its NGO partners such as BRAC and its associated consortia. However, our findings and analyses show that PSE will be an important tool for strengthening malaria surveillance and treatment networks as the country works towards elimination. This study distilled a voluminous literature to establish precise definitions for eight individual strategies for private sector engagement, weighing each based on the evidence provided in the literature and its applicability for Bangladesh. Based on this analysis, we recommend a phased, multi-modal model in which the first step combines RDT subsidization with rigorous provider training in high- and low-transmission as well as near-elimination settings. Subsequently we recommend that referral fees, which represented the most effective individual strategy studied, and possibly certification, infrastructure building, and demand-side intervention be introduced to better engage the private healthcare sector in malaria elimination efforts.

#### LIMITATIONS

Given the relative lack of evidence, this study did not analyze private sector engagement with regards to gender dynamics, which may limit its applicability. This study was also limited to sources in English, with the small exception oof a few sources in Bangla for which translations could be provided.

#### DATA SHARING STATEMENT

This study aims to provide full data transparency. A full list of search terms and sources included in the literature review can be found in the supplemental materials. All other inquiries for data sharing can be directed to the corresponding author.

#### ETHICS STATEMENT

No participant data was used in the formulation of this manuscript.

## Supporting information

Literature Search Terms and Results

## Data Availability

All data produced in the present study are available upon reasonable request to the authors

## Author Contributions

Ryan McArdle: Conceptualization, Investigation, Methodology, Visualization, Writing – Original Draft, Writing – Review and Editing

Ching Swe Phru: Project Administration, Supervision

Mohammad Sharif Hossain: Project Administration, Supervision

Mohammad Shafiul Alam: Conceptualization, Project Administration, Supervision, Writing – review and editing

Kasturi Haldar: Conceptualization, Funding Acquisition, Supervision, Project Administration, Visualization, Writing – Original Draft, Writing – Review and Editing

