## Supplementary material for "Bangladesh should engage the private sector for malaria elimination by 2030": Literature Search Terms and Results

**Supplemental Material for Opportunities for Private Sector Engagement in Malaria Elimination in Bangladesh**

**Full Literature Review Search Terms:**

- Private sector engagement reward healthcare
- Private sector engagement malaria
- Private sector engagement malaria Bangladesh
- Subsidization AND healthcare AND developing AND malaria
- Strategies private provider incentive healthcare
- Strategies private provider contracting malaria
- Strategies private provider contracting healthcare
- Strategies private provider contracting healthcare Bangladesh
- Social marketing healthcare private provider review
- Social marketing health evidence promotion AND developing OR LMIC
- Social franchising healthcare private
- Regulation AND private AND healthcare
- Regulate OR regulation OR regulatory AND disease reporting OR case reporting OR notifiable disease AND review
- RDT AND subsidization OR subsidize AND malaria
- RDT AND subsidization OR subsidize AND malaria AND review
- Public-private partnership OR public private partnership AND private sector OR private provider OR private sector engagement AND healthcare AND review
- Public awareness campaign systematic review healthcare
- Public awareness campaign disease evidence review AND Africa OR Asia OR Southeast Asia
- Private sector engagement AND health AND training AND review
- Private sector engagement AND case reporting AND reward OR reimbursement OR honorarium OR pay OR cash
- Private provider AND self-regulation OR self regulation AND evidence AND health
- Pay for performance OR p4p AND review
- Notifiable disease AND reporting AND reward OR incentive OR pay
- Health AND LMIC AND review AND patient AND demand-side interventions
- Health AND LMIC AND review AND community mobilization
- AMFm systematic review

**Full List of Articles Included in Study: (Full search results available upon request)**

1. World malaria report 2022. Geneva: World Health Organization; 2022.
2. National Malaria Elimination Programme. NATIONAL STRATEGIC PLAN FOR MALARIA ELIMINATION IN BANGLADESH: 2021-2025 2021; <https://apmen.org/sites/default/files/all_resources/National%20Strategic%20Plan_Malaria%20Elimination_Bangladesh_2021-2025.pdf>.
3. Matin, Mohammad Abdul, et al. "Precarity at the margins of malaria control in the Chittagong Hill Tracts in Bangladesh: a mixed-methods study." *Pathogens* 9.10 (2020): 840.
4. Galagan, Sean R., et al. "The practice of jhum cultivation and its relationship to Plasmodium falciparum infection in the Chittagong Hill Districts of Bangladesh." The American journal of tropical medicine and hygiene 91.2 (2014): 374.
5. Sinha I, Sayeed AA, Uddin D, et al. Mapping the travel patterns of people with malaria in Bangladesh*. BMC Medicine* 2020; 18: 45. DOI:10.1186/s12916-020-1512-5.
6. Haldar K, Alam MS, Koepfli C, et al. Bangladesh in the era of malaria elimination*. Trends in parasitology*. DOI:10.1016/j.pt.2023.06.009.
7. Potter R, Tesfazghi K, Poyer S, et al. Private Sector Contributions to National Malaria Surveillance Systems in Elimination Settings: Lessons Learned from Cambodia, Lao PDR, Myanmar, and Vietnam*. The American journal of tropical medicine and hygiene* 2023; 108: 14-23. DOI:10.4269/ajtmh.22-0147.
8. Directorate General of Health Services MoHaFW, Government of the People’s Republic of Bangladesh. Health Bulletin 2019. 2020.
9. Government of the People's Republic of Bangladesh. Communicable Diseases (Prevention, Control and Eradication) Act, 2018
   (Act No. 61 of 2018) 2018;.
10. Ley, Benedikt, et al. "Barriers to routine G6PD testing prior to treatment with primaquine." Malaria journal 16.1 (2017): 1-10.
11. Engel, Nora, et al. "Implementing radical cure diagnostics for malaria: user perspectives on G6PD testing in Bangladesh." Malaria journal 20.1 (2021): 1-12.
12. Bhuiya, A. "Health for the rural masses: Insights from Chakaria: ICDDR." (2009).
13. Cotter, Chris, et al. "The changing epidemiology of malaria elimination: new strategies for new challenges." The Lancet382.9895 (2013): 900-911.
14. Hopkins, Donald R., and Ernesto Ruiz-Tiben. "Surveillance for dracunculiasis, 1981–1991." *MORBIDITY AND MORTALITY WEEKLY REPORT: CDC Surveillance Summaries* (1992): 1-13.
15. Foster, Stanley O., and Abdullahi Deria. "Smallpox eradication in Somali nomadic encampments: The search for a culturally acceptable method of case detection, case isolation, and outbreak control." *Medical Anthropology* 7.2 (1983): 19-25.
16. World Health Organization. "Preparing for certification of malaria elimination." (2020).
17. Husain, Mohammed, et al. "Disease surveillance system of Bangladesh: combating public health emergencies." Online Journal of Public Health Informatics 11.1 (2019).
18. Bennett, Adam, et al. "Engaging the private sector in malaria surveillance: a review of strategies and recommendations for elimination settings." *Malaria journal* 16.1 (2017): 1-19.
19. Montagu, Dominic, et al. "Recent trends in working with the private sector to improve basic healthcare: a review of evidence and interventions." *Health Policy and Planning* 31.8 (2016): 1117-1132.
20. Shah, Nirali M., William R. Brieger, and David H. Peters. "Can interventions improve health services from informal private providers in low and middle-income countries? A comprehensive review of the literature." *Health Policy and Planning* 26.4 (2011): 275-287.
21. Onoka CA, Onwujekwe OE, Hanson K, et al. Sub-optimal delivery of intermittent preventive treatment for malaria in pregnancy in Nigeria: influence of provider factors*. Malaria Journal* 2012; 11: 317. DOI:10.1186/1475-2875-11-317.
22. Waters, Hugh, Laurel Hatt, and David Peters. "Working with the private sector for child health." *Health policy and planning*18.2 (2003): 127-137.
23. Adams, Alayne Mary, et al. "Exploratory qualitative study to understand the underlying motivations and strategies of the private for-profit healthcare sector in urban Bangladesh." *BMJ open* 9.7 (2019): e026586.
24. Ali, M. S. "Innovative health management–RahimYar Khan District, Islamabad." *Islamabad: CIDA Devolution Support Project* (2005).
25. Schwartz, J. and Bhushan, I., 2004, 'Cambodia: Using Contracting to Reduce Inequity in Primary Health Care Delivery', HNP Discussion paper, Reaching the Poor Programme Paper no. 3, World Bank, Washington DC
26. Bloom, Erik, et al. "Contracting for health: evidence from Cambodia." *Brookings Institution* (2006).
27. Marek, Tonia, et al. "Successful contracting of prevention services: fighting malnutrition in Senegal and Madagascar." *Health Policy and planning* 14.4 (1999): 382-389.
28. Cristia, Julian, Ariadna García Prado, and Cecilia Peluffo. "The impact of contracting in and contracting out basic health services: the Guatemalan experience." *World Development* 70 (2015): 215-227.
29. Hiwat, Hélène, et al. "Novel strategies lead to pre-elimination of malaria in previously high-risk areas in Suriname, South America." *Malaria Journal* 11.1 (2012): 1-12.
30. Heard, Anna, Dhiraj Kumar Nath, and Benjamin Loevinsohn. "Contracting urban primary healthcare services in Bangladesh–effect on use, efficiency, equity and quality of care." *Tropical Medicine & International Health* 18.7 (2013): 861-870.
31. Loevinsohn, Benjamin, and April Harding. "Buying results? Contracting for health service delivery in developing countries." *The Lancet* 366.9486 (2005): 676-681.
32. Lagarde, Mylene, and Natasha Palmer. "The impact of contracting out on health outcomes and use of health services in low and middle‐income countries." *Cochrane Database of Systematic Reviews* 4 (2009).
33. Odendaal, Willem A., et al. "Contracting out to improve the use of clinical health services and health outcomes in low‐and middle‐income countries." *Cochrane Database of Systematic Reviews* 4 (2018).
34. Liu, Xingzhu, David R. Hotchkiss, and Sujata Bose. "The effectiveness of contracting-out primary health care services in developing countries: a review of the evidence." *Health policy and planning* 23.1 (2008): 1-13.
35. “Operations Evaluation Department. 2005. Maintaining Momentum to 2015 : An Impact Evaluation of Interventions to Improve Maternal and Child Health and Nutrition in Bangladesh”. Washington, DC: World Bank.
36. Peters, David H., Gita G. Mirchandani, and Peter M. Hansen. "Strategies for engaging the private sector in sexual and reproductive health: how effective are they?." *Health policy and planning* 19.suppl_1 (2004): i5-i21.
37. Menya, Diana, et al. "Using pay for performance incentives (P4P) to improve management of suspected malaria fevers in rural Kenya: a cluster randomized controlled trial." *BMC medicine* 13.1 (2015): 1-13.
38. Chowdhury AMR, Ahmed YH, Islam K, et al. *50 years of Bangladesh : Advances in Health*. Dhaka: University Press Ltd, 2023.
39. Friedman, Jed, Ashis Das, and Ronald Mutasa. "Rewarding Provider Performance to Improve Quality and Coverage of Maternal and Child Health Outcomes: Zimbabwe Results-Based Financing Pilot Program." *Washington, DC: The World Bank* (2016).
40. Zang, Omer, et al. "Impact of performance-based financing on health-care quality and utilization in urban areas of Cameroon." *African health monitor* (2015).
41. De Walque, Damien, et al. "Looking into the performance-based financing black box: evidence from an impact evaluation in the health sector in Cameroon." *World Bank Policy Research Working Paper* 8162 (2017).
42. Witter S, Fretheim A, Kessy FL, et al. Paying for performance to improve the delivery of health interventions in low‐ and middle‐income countries*. Cochrane database of systematic reviews* 2012; 2013: CD007899. DOI:10.1002/14651858.CD007899.pub2
43. Diaconu K, Witter S, Binyaruka P, et al. Appraising pay-for-performance in healthcare in low- and middle-income countries through systematic reviews: reflections from two teams*. Cochrane database of systematic reviews* 2022; 5: ED000157. DOI:10.1002/14651858.ED000157.
44. Kondo, Karli K., et al. "Implementation processes and pay for performance in healthcare: a systematic review." *Journal of general internal medicine* 31.1 (2016): 61-69.
45. Van Herck, Pieter, et al. "Systematic review: effects, design choices, and context of pay-for-performance in health care." *BMC health services research* 10.1 (2010): 1-13.
46. Eijkenaar, Frank, et al. "Effects of pay for performance in health care: a systematic review of systematic reviews." *Health policy* 110.2-3 (2013): 115-130.
47. Doyle, Timothy J., M. Kathleen Glynn, and Samuel L. Groseclose. "Completeness of notifiable infectious disease reporting in the United States: an analytical literature review." *American journal of epidemiology* 155.9 (2002): 866-874.
48. Tan, Hsiu-Fen, et al. "Private doctors' practices, knowledge, and attitude to reporting of communicable diseases: a national survey in Taiwan." *BMC infectious diseases* 9.1 (2009): 1-8.
49. Schramm, Margaret M., Richard L. Vogt, and Margaret Mamolen. "The surveillance of communicable disease in Vermont: who reports?." *Public Health Reports* 106.1 (1991): 95.
50. Revere, Debra, et al. "Notifiable condition reporting practices: implications for public health agency participation in a health information exchange." *BMC public health* 17.1 (2017): 1-12.
51. Tan, Hsiu-Fen, et al. "Evaluation of the National Notifiable Disease Surveillance System in Taiwan: an example of varicella reporting." *Vaccine* 25.14 (2007): 2630-2633.
52. Cao, Jun, et al. "Communicating and monitoring surveillance and response activities for malaria elimination: China's “1-3-7” strategy." *PLoS medicine* 11.5 (2014): e1001642.
53. Chiang, Chen Yuan, et al. "The impact of national health insurance on the notification of tuberculosis in Taiwan." *The International Journal of Tuberculosis and Lung Disease* 6.11 (2002): 974-979.
54. Lo, Hsiu-Yun, et al. "Completeness and timeliness of tuberculosis notification in Taiwan." *BMC public health* 11.1 (2011): 1-9.
55. Vo, Luan Nguyen Quang, et al. "Enhanced private sector engagement for tuberculosis diagnosis and reporting through an intermediary agency in Ho Chi Minh City, Viet Nam." *Tropical medicine and infectious disease* 5.3 (2020): 143.
56. Senyonjo, Laura, et al. "Lessons learned for surveillance strategies for trachoma elimination as a public health problem, from the evaluation of approaches utilised by Guinea worm and onchocerciasis programmes: A literature review." *PLoS neglected tropical diseases* 15.1 (2021): e0009082.
57. World Health Organization. *The global eradication of smallpox: final report of the Global Commission for the Certification of Smallpox Eradication, Geneva, December 1979*. World Health Organization, 1980.
58. Kambo, Indra P., et al. "Use of traditional medical practitioners to deliver family planning services in Uttar Pradesh." *Studies in Family Planning* (1994): 32-40.
59. Garcia, Patricia, et al. "Training pharmacy workers in recognition, management, and prevention of STDs: district-randomized controlled trial." *Bulletin of the World Health Organization* 81.11 (2003): 806-814.
60. AlMossawi, Hala Jassim, et al. "Universal Health Coverage-Inclusion of TB in National Health Insurance Programs and Recommendations for Expansion of Coverage of TB Services in LMICs." *Journal of Lung Health and Diseases* 3.3 (2019).
61. Wells, William A., Mukund Uplekar, and Madhukar Pai. "Achieving systemic and scalable private sector engagement in tuberculosis care and prevention in Asia." *PLoS medicine*12.6 (2015): e1001842.
62. Enserink, Martin. "Malaria drugs, the Coca-Cola way." *Science*322.5905 (2008): 1174-1174.
63. Matowe, Lloyd, and Olusoji Adeyi. "The quest for universal access to effective malaria treatment: how can the AMFm contribute?." Malaria Journal 9.1 (2010): 1-5.
64. Lussiana, Cristina. "Towards subsidized malaria rapid diagnostic tests. Lessons learned from programmes to subsidise artemisinin-based combination therapies in the private sector: a review." *Health policy and planning* 31.7 (2016): 928-939.
65. Frost, L., and M. R. Reich. "Translating an idea into a policy:“Saving lives and buying time” for antimalarial medicines." *Harvard: Harvard School of Public Health* (2009).
66. Kamal-Yanni, Mohga. "Salt, sugar, and malaria pills: how the Affordable Medicine Facility–malaria endangers public health." (2012).
67. Goodman, C., et al. "Independent evaluation of phase 1 of the Affordable medicines Facility-malaria (AMFm), Multi-Country independent evaluation final report." (2012).
68. Morris, Alexandra, et al. "Price subsidies increase the use of private sector ACTs: evidence from a systematic review." *Health policy and planning* 30.3 (2015): 397-405.
69. Ikwuobe, John O., et al. "The impact of rapid malaria diagnostic tests upon anti-malarial sales in community pharmacies in Gwagwalada, Nigeria." *Malaria Journal* 12.1 (2013): 1-8.
70. Ansah, Evelyn K., et al. "The impact of providing rapid diagnostic malaria tests on fever management in the private retail sector in Ghana: a cluster randomized trial." *bmj* 350 (2015).
71. Cohen, Jessica, et al. "Feasibility of distributing rapid diagnostic tests for malaria in the retail sector: evidence from an implementation study in Uganda." *PloS one* 7.11 (2012): e48296.
72. Cohen, Dupas, P., & Schaner, S. (2015). Price Subsidies, Diagnostic Tests, and Targeting of Malaria Treatment: Evidence from a Randomized Controlled Trial. *The American Economic Review*, *105*(2), 609–645.
73. Maloney, Kathleen, et al. "Expanding access to parasite-based malaria diagnosis through retail drug shops in Tanzania: evidence from a randomized trial and implications for treatment." *Malaria journal* 16.1 (2017): 1-10.
74. Laktabai, Jeremiah, et al. "Subsidise the test, the treatment or both? Results of an individually randomised controlled trial of the management of suspected malaria fevers in the retail sector in western Kenya." *BMJ global health* 5.11 (2020): e003378.
75. Visser, Theodoor, et al. "Introducing malaria rapid diagnostic tests in private medicine retail outlets: a systematic literature review." *PloS one* 12.3 (2017): e0173093.
76. Hanson, Kara, and Catherine Goodman. "Testing times: trends in availability, price, and market share of malaria diagnostics in the public and private healthcare sector across eight sub-Saharan African countries from 2009 to 2015." *Malaria journal*16.1 (2017): 1-16.
77. O'Connell, Kathryn, et al. "Using and joining a franchised private sector provider network in Myanmar." *PLoS One* 6.12 (2011): e28364.
78. Bishai, David M., et al. "A social franchising to improve quality and access in private health care in developing countries." (2008): 184-197.
79. Patouillard, Edith, et al. "Can working with the private for-profit sector improve utilization of quality health services by the poor? A systematic review of the literature." *International journal for equity in health* 6.1 (2007): 1-11.
80. Nijmeijer, Karlijn J., Isabelle N. Fabbricotti, and Robbert Huijsman. "Making franchising work: A framework based on a systematic review." *International Journal of Management Reviews* 16.1 (2014): 62-83.
81. Koehlmoos, T. P., et al. "Social franchising evaluations: a scoping review. London: EPPI-Centre." *Social Science Research Unit, Institute of Education, University of London*(2011): 1-58.
82. Cumberland, Denise M., and Benjamin C. Litalien. "Social franchising: A systematic review." *Journal of Marketing Channels* 25.3 (2018): 137-156.
83. Beyeler, Naomi, Anna York De La Cruz, and Dominic Montagu. "The impact of clinical social franchising on health services in low-and middle-income countries: a systematic review." *PloS one* 8.4 (2013): e60669.
84. Montagu, Dominic, and Catherine Goodman. "Prohibit, constrain, encourage, or purchase: how should we engage with the private health-care sector?." *The Lancet* 388.10044 (2016): 613-621.
85. Ngo, Anh D., et al. "The impact of social franchising on the use of reproductive health and family planning services at public commune health stations in Vietnam." *BMC Health Services Research* 10.1 (2010): 1-8.
86. Haemmerli, Manon, et al. "How equitable is social franchising? Case studies of three maternal healthcare franchises in Uganda and India." *Health policy and planning* 33.3 (2018): 411-419.
87. Morgan, Rosemary, Tim Ensor, and Hugh Waters. "Performance of private sector health care: implications for universal health coverage." *The Lancet* 388.10044 (2016): 606-612.
88. World Health Organization. (‎2003)‎. Quality and accreditation in health care services : a global review. World Health Organization.
89. Morgan, Rosemary, and Tim Ensor. "The regulation of private hospitals in Asia." *The International Journal of Health Planning and Management* 31.1 (2016): 49-64.
90. Tangcharoensathien, V., et al. "Regulation of health service delivery in private sector: challenges and opportunities." *The Rockefeller Foundation Technical Partner Paper* 9 (2008).
91. Alkhenizan, Abdullah, and Charles Shaw. "Impact of accreditation on the quality of healthcare services: a systematic review of the literature." *Annals of Saudi medicine*31.4 (2011): 407-416.
92. Chakravarthi, Indira. "Regulation of private health care providers in India: Current status, future directions." *Indian Journal of Public Administration* 64.4 (2018): 587-598.
93. Stenson, Bo, et al. "Private pharmacy practice and regulation: a randomized trial in Lao PDR." *International journal of technology assessment in health care* 17.4 (2001): 579-589.
94. Vijaykadga, Saowanit, et al. "Strengthening of national capacity in implementation of antimalarial drug quality assurance in Thailand." *Southeast Asian journal of tropical medicine and public health* 37 (2006): 5.
95. MOHFW, Government of India. "Situational Analyses: Backdrop to the National Health Policy - 2017." (2017): 31.
96. Sheikh, Kabir, Prasanna S. Saligram, and Krishna Hort. "What explains regulatory failure? Analysing the architecture of health care regulation in two Indian states." *Health Policy and Planning* 30.1 (2015): 39-55.
97. Benson, Frew Gerald, Jonathan Levin, and Laetitia Charmaine Rispel. "Health care providers’ compliance with the notifiable diseases surveillance system in South Africa." *PloS one* 13.4 (2018): e0195194.
98. Friedman, S. M., et al. "ARCHIVED-Suboptimal reporting of notifiable diseases in Canadian emergency departments: A survey of emergency physician knowledge, practices, and perceived barriers." *Canada Communicable Disease Report*32.17 (2006).
99. Dixon, Brian E., et al. "Completeness and timeliness of notifiable disease reporting: a comparison of laboratory and provider reports submitted to a large county health department." *BMC medical informatics and decision making*17.1 (2017): 1-8.
100. Wafula, Francis, et al. "Protecting the public or setting the bar too high? Understanding the causes and consequences of regulatory actions of front-line regulators and specialized drug shop operators in Kenya." *Social science & medicine* 97 (2013): 220-227.
101. Harding, April, and Alexander S. Preker, eds. *Private participation in health services*. Vol. 434. World Bank Publications, 2003.
102. Ayres, Ian, and John Braithwaite. "Partial-industry regulation: A monopsony standard for consumer protection." *Calif. L. Rev.*80 (1992): 13.
103. World Health Organization. "Public policy and franchising reproductive health: current evidence and future directions Guidance from a technical consultation meeting." (2007).
104. McPake, Barbara, and Kara Hanson. "Managing the public–private mix to achieve universal health coverage." *The Lancet*388.10044 (2016): 622-630.
105. Ensor, Tim, and Sabine Weinzierl. "Regulating health care in low-and middle-income countries: Broadening the policy response in resource constrained environments." *Social Science & Medicine* 65.2 (2007): 355-366.
106. World Health Organization. *The world health report 2006: working together for health*. World Health Organization, 2006.
107. Bennett, Sara, Ellias E. Ngalande-Banda, and World Health Organization. "Public and private roles in health: a review and analysis of experience in Sub-Saharan Africa." (1994).
108. Chandler, Clare IR, et al. "Introducing malaria rapid diagnostic tests at registered drug shops in Uganda: limitations of diagnostic testing in the reality of diagnosis." *Social science & medicine* 72.6 (2011): 937-944.
109. Argaw, Mesele Damte, Thandisizwe Redford Mavundla, and Kassa Daka Gidebo. "Community and healthcare providers’ perceptions of quality of private sector outpatient malaria care in North-western Ethiopia: a qualitative study." *Malaria Journal*20.1 (2021): 1-11.
110. Palafox, Benjamin, et al. "Understanding private sector antimalarial distribution chains: a cross-sectional mixed methods study in six malaria-endemic countries." *PloS one* 9.4 (2014): e93763.
111. Déglise, Carole, L. Suzanne Suggs, and Peter Odermatt. "Short message service (SMS) applications for disease prevention in developing countries." *Journal of medical Internet research* 14.1 (2012): e1823.
112. Zabih, Weeda, et al. "Interventions to improve early detection of childhood cancer in low‐and middle‐income countries: a systematic review." *Pediatric Blood & Cancer* 67.12 (2020): e28761.
113. Nam, Yoon-Young, et al. "Creating public awareness in Asia of depression as treatable and suicide as preventable." *Suicide and suicide prevention in Asia* (2008): 31-38.
114. Borawska, Anna. "The role of public awareness campaigns in sustainable development." *Economic and Environmental Studies* 17.44 (2017): 865-877.
115. Naugle, Danielle A., and Robert C. Hornik. "Systematic review of the effectiveness of mass media interventions for child survival in low-and middle-income countries." *Journal of health communication* 19.sup1 (2014): 190-215.
116. Schiavo, Renata, May May Leung, and Mason Brown. "Communicating risk and promoting disease mitigation measures in epidemics and emerging disease settings." *Pathogens and Global Health* 108.2 (2014): 76-94.
117. Hurst, Taylor E., et al. "Demand-side interventions for maternal care: evidence of more use, not better outcomes." *BMC pregnancy and childbirth* 15.1 (2015): 1-14.
118. Hasanov, E., et al. "Assessing the impact of public education on a preventable zoonotic disease: rabies." *Epidemiology & Infection* 146.2 (2018): 227-235.
119. Cao, Man, et al. "Impact of a health campaign on Chinese public awareness of stroke: evidence from internet search data." *BMJ open* 11.12 (2021): e054463.
120. Omedo, Martin, et al. "The effect of a health communication campaign on compliance with mass drug administration for schistosomiasis control in western Kenya—the SCORE project." *The American journal of tropical medicine and hygiene* 91.5 (2014): 982.
121. Graziose, Matthew M., et al. "Systematic review of the design, implementation and effectiveness of mass media and nutrition education interventions for infant and young child feeding." *Public health nutrition* 21.2 (2018): 273-287.
122. Schliemann, Désirée, et al. "Impact of a mass media campaign on breast cancer symptoms awareness and screening uptake in Malaysia: Findings from a quasi-experimental study." *BMJ open* 10.8 (2020): e036503.
123. Dada, Sara, et al. "Community mobilization to strengthen support for appropriate and timely use of antenatal and postnatal care: A review of reviews." *Journal of Global Health*11 (2021).
124. Beck, Dana C., Michelle L. Munro-Kramer, and Jody R. Lori. "A scoping review on community mobilisation for maternal and child health in sub-Saharan Africa: Impact on empowerment." *Global public health* 14.3 (2019): 375-395.
125. Tiruneh, Gizachew Tadele, Chalachew Bekele Shiferaw, and Alemayehu Worku. "Effectiveness and cost-effectiveness of home-based postpartum care on neonatal mortality and exclusive breastfeeding practice in low-and-middle-income countries: a systematic review and meta-analysis." *BMC pregnancy and childbirth* 19.1 (2019): 1-19.
126. Kiragu, Michael, et al. "Does capacity development increase demand for health services and rights among key populations affected by HIV? A systematic review of evidence from low and middle-income countries." *AIDS and Behavior* 24.8 (2020): 2268-2281.
127. Safieh, Jacqueline, et al. "Reported evidence on the effectiveness of mass media interventions in increasing knowledge and use of family planning in low and middle-income countries: a systematic mixed methods review." *Journal of global health* 9.2 (2019).
128. Ezennia, Ifeoma, and Obinna Ekwunife. "COST-BENEFIT ANALYSIS OF MALARIA RAPID DIAGNOSTIC TEST IN ENUGU METROPOLIS, NIGERIA: THE PERSPECTIVE OF THE COMMUNITY PHARMACY PRACTITIONER." BMJ Global Health 2.Suppl 2 (2017).
129. Stead, Martine, et al. "A systematic review of social marketing effectiveness." *Health education* 107.2 (2007): 126-191.
130. Evans, W. Douglas, et al. "Systematic review of public health branding." *Journal of health communication* 13.8 (2008): 721-741.
131. Krishnaratne, Shari, et al. "Interventions to strengthen the HIV prevention cascade: a systematic review of reviews." *The lancet HIV* 3.7 (2016): e307-e317.
132. Lassi, Zohra S., et al. "Strategies for improving health care seeking for maternal and newborn illnesses in low-and middle-income countries: a systematic review and meta-analysis." *Global health action* 9.1 (2016): 31408.
133. Brown, Morgan, et al. "Motivation to test, treat, and report malaria cases: a quantitative assessment among private sector providers in the Greater Mekong Subregion." *Malaria Journal* 21.1 (2022): 1-17.
134. World Health Organization. "Towards better engagement of the private sector in health service delivery: a review of approaches to private sector engagement in Africa." (2022).
135. Sadeghi, Ahmad, et al. "Experiences of selected countries in the use of public-private partnership in hospital services provision." *J Pak Med Assoc* 66.11 (2016): 1401-6.
136. Jones, Robert T., et al. "The role of the private sector in supporting malaria control in resource development settings." *The Journal of Infectious Diseases* 222.Supplement_8 (2020): S701-S708.
137. Public-Private Partnership Legal Resource Center. “About Public-Private Partnerships”. *The World Bank*. 21 June 2022
138. Spielman, Andrew, et al. "Industrial anti-malaria policies." *Center for International Development and Harvard School of Public Health, Cambridge and Boston (prepared for the World Economic Forum 2002)*. 2002.
139. Argaw, Mesele D., et al. "Improved malaria case management in formal private sector through public private partnership in Ethiopia: retrospective descriptive study." *Malaria journal* 15.1 (2016): 1-11.
140. Billingsley, Peter F., et al. "The Equatoguinean malaria vaccine initiative: from the launching of a clinical research platform to malaria elimination planning in Central West Africa." *The American journal of tropical medicine and hygiene* 103.3 (2020): 947.
141. Fernando, Deepika, et al. "Use of a public-private partnership in malaria elimination efforts in Sri Lanka; a case study." *BMC Health Services Research* 18.1 (2018): 1-10.
142. Ferdiana, Astri, et al. "Finding the right balance: implementation of public–private partnership in artemisinin-based combination therapy provision in Manokwari, Indonesia." *Journal of Pharmaceutical Policy and Practice*14.1 (2021): 1-12.
143. Yadav, Vikas, et al. "Facilitators and barriers to participation of private sector health facilities in government-led schemes for maternity services in India: a qualitative study." *BMJ open* 7.6 (2017): e017092.
144. Smith, Lucy A., et al. "Provider practice and user behavior interventions to improve prompt and effective treatment of malaria: do we know what works?." *The American journal of tropical medicine and hygiene* 80.3 (2009): 326-335.
145. Ghouth, Abdulla Salim Bin. "Availability and prescription practice of anti-malaria drugs in the private health sector in Yemen." *The Journal of Infection in Developing Countries* 7.05 (2013): 404-412.
146. Esena, Reuben K., and Francis M. Djangmah. "Provider Training on the use of AMFm ACTs in Private Drug Outlets in Ghana: a case study on the impact of malaria services in urban areas." *Journal of Scientific and Innovative Research*2.3 (2013): 512-517.
147. Argaw, Mesele Damte. "Knowledge and practice on malaria diagnosis and treatment among healthcare providers working in private health facilities in Ethiopia." *Malar World J* 6.10 (2015): 1-8.
148. Rao, V. Bhargavi, David Schellenberg, and Azra C. Ghani. "Overcoming health systems barriers to successful malaria treatment." *Trends in Parasitology* 29.4 (2013): 164-180.
149. Hill, Jenny, et al. "Women's access and provider practices for the case management of malaria during pregnancy: a systematic review and meta-analysis." *PLoS medicine* 11.8 (2014): e1001688.
150. Kamal-Yanni, Mohga M., Julien Potet, and Philippa M. Saunders. "Scaling-up malaria treatment: a review of the performance of different providers." *Malaria journal* 11.1 (2012): 1-10.
151. Ratsimbasoa, Arsène, et al. "Management of uncomplicated malaria in febrile under five-year-old children by community health workers in Madagascar: reliability of malaria rapid diagnostic tests." *Malaria Journal* 11.1 (2012): 1-8.
152. Bosch–Capblanch, Xavier, and Claudine Marceau. "Training, supervision and quality of care in selected integrated community case management (iCCM) programmes: a scoping review of programmatic evidence." *Journal of Global Health*4.2 (2014).
153. Smith Paintain, Lucy, et al. "Intermittent screening and treatment versus intermittent preventive treatment of malaria in pregnancy: provider knowledge and acceptability." *PloS one*6.8 (2011): e24035.
154. Willey, Barbara A., et al. "Communicating the AMFm message: exploring the effect of communication and training interventions on private for-profit provider awareness and knowledge related to a multi-country anti-malarial subsidy intervention." *Malaria journal* 13.1 (2014): 1-10.
155. Tetteh, Mary, et al. "Impact of malaria diagnostic refresher training programme on competencies and skills in malaria diagnosis among medical laboratory professionals: evidence from Ghana 2015–2019." *Malaria Journal* 20.1 (2021): 1-12.
156. Wasunna, Beatrice, et al. "Health worker performance in the management of paediatric fevers following in-service training and exposure to job aids in Kenya." *Malaria Journal* 9.1 (2010): 1-7.
157. Gindola, Yamlak, et al. "Adherence to national malaria clinical management and testing guidelines in selected private clinics of Gambela Town, Gambela Region, Ethiopia: a mixed method study." *Malaria Journal* 21.1 (2022): 1-8.
158. Ofori-Adjei, David, and Daniel K. Arhinful. "Effect of training on the clinical management of malaria by medical assistants in Ghana." *Social science & medicine* 42.8 (1996): 1169-1176.
159. Hamer, Davidson H., et al. "Improved diagnostic testing and malaria treatment practices in Zambia." *Jama* 297.20 (2007): 2227-2231.
160. Dräger, Sigrid, Gulin Gedik, and Mario R. Dal Poz. "Health workforce issues and the Global Fund to fight AIDS, Tuberculosis and Malaria: an analytical review." *Human Resources for Health* 4.1 (2006): 1-12.
161. Wasunna, Beatrice, et al. "Why don't health workers prescribe ACT? A qualitative study of factors affecting the prescription of artemether-lumefantrine." *Malaria Journal* 7.1 (2008): 1-9.
162. Rahman R. The State, the Private Health Care Sector and Regulation in Bangladesh*. Asia Pacific journal of public administration* 2007; 29: 191-206. DOI:10.1080/23276665.2007.10779334.
163. Zurovac, Dejan, Ambrose O. Talisuna, and Robert W. Snow. "Mobile phone text messaging: tool for malaria control in Africa." *PLoS medicine* 9.2 (2012): e1001176.
164. Kangwana, Beth P., et al. "The effect of an anti-malarial subsidy programme on the quality of service provision of artemisinin-based combination therapy in Kenya: a cluster-randomized, controlled trial." *Malaria journal* 12.1 (2013): 1-13.
165. Kangwana, Beth P., et al. "The impact of retail-sector delivery of artemether–lumefantrine on malaria treatment of children under five in Kenya: a cluster randomized controlled trial." *PLoS medicine* 8.5 (2011): e1000437.
166. Kedenge, Sarah V., et al. "Understanding the impact of subsidizing artemisinin-based combination therapies (ACTs) in the retail sector–results from focus group discussions in rural Kenya." *PloS one* 8.1 (2013): e54371.
167. Aung, Tin, et al. "Improving uptake and use of malaria rapid diagnostic tests in the context of artemisinin drug resistance containment in eastern Myanmar: an evaluation of incentive schemes among informal private healthcare providers." *Malaria Journal* 14.1 (2015): 1-9.
168. Cohen JM. “Remarkable solutions to impossible problems”: lessons for malaria from the eradication of smallpox. *Malaria journal*. 2019 Dec;18:1-6.
169. Seidman G, Atun R. Do changes to supply chains and procurement processes yield cost savings and improve availability of pharmaceuticals, vaccines or health products? A systematic review of evidence from low-income and middle-income countries. *BMJ global health*. 2017 Apr 1;2(2):e000243.
170. Koduah A, Baatiema L, de Chavez AC, Danso-Appiah A, Kretchy IA, Agyepong IA, King N, Ensor T, Mirzoev T. Implementation of medicines pricing policies in sub-Saharan Africa: systematic review. Systematic reviews. 2022 Dec 1;11(1):257.
171. Salam RA, Das JK, Lassi ZS, Bhutta ZA. Impact of community-based interventions for the prevention and control of malaria on intervention coverage and health outcomes for the prevention and control of malaria. Infectious diseases of poverty. 2014 Dec;3:1-5.
172. Awor P, Miller J, Peterson S. Systematic literature review of integrated community case management and the private sector in Africa: relevant experiences and potential next steps. Journal of global health. 2014 Dec;4(2).
173. Silverman R, Rosen D, Regan L, Vernon J, Yadav P. Malaria case management in the private sector in Africa: a call for action to identify sustainable solutions. Center for Global Development. 2021 Aug.
174. Ilesanmi O, Oton E, Afolabi A. Community Engagement in the Prevention and Control of Lassa Fever in Africa: A Systematic Review. Infection Epidemiology and Microbiology. 2021 May 10;7(2):187-96.
175. George AS, Scott K, Mehra V, Sriram V. Synergies, strengths and challenges: findings on community capability from a systematic health systems research literature review. BMC health services research. 2016 Nov;16:47-59.
176. An evidence map of social, behavioural and community engagement interventions for reproductive, maternal, newborn and child health. Geneva: World Health Organization; 2017. https://iris.who.int/bitstream/handle/10665/259399/9789240697263-eng.pdf
177. Parmar D, Banerjee A. How do supply-and demand-side interventions influence equity in healthcare utilisation? Evidence from maternal healthcare in Senegal. Social Science & Medicine. 2019 Nov 1;241:112582.
178. James N, Lawson K, Acharya Y. Evidence on result-based financing in maternal and child health in low-and middle-income countries: a systematic review. Global health research and policy. 2020 Dec;5:1-5.
179. Gandhi S, Ramesh S, Dash U, Babu SM. A systematic review of Demand-based & Supply-based Interventions on continuum of maternal and child healthcare in south Asian countries. Journal of Public Health. 2021 Aug;29:857-70.
180. Rivera M, Miller K, Gomanie NN, Mehta K. A Narrative Review of Demand Generation Strategies For Family Planning in Low-and Middle-Income Countries. In2022 IEEE Global Humanitarian Technology Conference (GHTC) 2022 Sep 8 (pp. 341-348). IEEE.
181. Witter, S. Pay for performance for strengthening delivery of sexual and reproductive health services in low- and middle-income countries Evidence synthesis paper. The World Bank. 2013. https://eresearch.qmu.ac.uk/handle/20.500.12289/3283
182. Cuevas C, Batura N, Wulandari LP, Khan M, Wiseman V. Improving antibiotic use through behaviour change: a systematic review of interventions evaluated in low-and middle-income countries. Health Policy and Planning. 2021 Jun 1;36(5):754-73.
183. Neelsen, Sven, de Walque, Damien, Friedman, Jed, and Wagstaff, Adam. Financial Incentives to Increase Utilization of Reproductive, Maternal, and Child Health Services in Low- and Middle-Income Countries: A Systematic Review and Meta-Analysis. 2021. Policy Research Working Paper; No. 9793. World Bank, Washington, DC. http://hdl.handle.net/10986/36344
184. Jain M, Shisler S, Lane C, Bagai A, Brown E, Engelbert M, Vardy Y, Eyers J, Leon DA, Parsekar SS. Use of community engagement interventions to improve child immunisation in low‐and middle‐income countries: A systematic review and meta‐analysis. Campbell Systematic Reviews. 2022 Sep;18(3):e1253.
185. Cross EL, Tolfree R, Kipping R. Systematic review of public-targeted communication interventions to improve antibiotic use. Journal of Antimicrobial Chemotherapy. 2017 Apr 1;72(4):975-87.
186. Higgs ES, Goldberg AB, Labrique AB, Cook SH, Schmid C, Cole CF, Obregón RA. Understanding the role of mHealth and other media interventions for behavior change to enhance child survival and development in low-and middle-income countries: an evidence review. Journal of health communication. 2014 May 6;19(sup1):164-89.
187. Stead M, Angus K, Langley T, Katikireddi SV, Hinds K, Hilton S, Lewis S, Thomas J, Campbell M, Young B, Bauld L. Mass media for public health messages: reviews of the evidence. Public Health Research. 2018 Jun 15.
188. Janati A, Hosseiny M, Gouya MM, Moradi G, Ghaderi E. Communicable disease reporting systems in the world: a systematic review article. Iranian journal of public health. 2015 Nov;44(11):1453.
189. Ng’etich AK, Voyi K, Kirinyet RC, Mutero CM. A systematic review on improving implementation of the revitalised integrated disease surveillance and response system in the African region: a health workers’ perspective. PLoS One. 2021 Mar 19;16(3):e0248998.
190. Emmert M, Eijkenaar F, Kemter H, Esslinger AS, Schöffski O. Economic evaluation of pay-for-performance in health care: a systematic review. The European Journal of Health Economics. 2012 Dec;13:755-67.
191. Kovacs RJ, Powell-Jackson T, Kristensen SR, Singh N, Borghi J. How are pay-for-performance schemes in healthcare designed in low-and middle-income countries? Typology and systematic literature review. BMC Health Services Research. 2020 Dec;20:1-4.
192. Singh NS, Kovacs RJ, Cassidy R, Kristensen SR, Borghi J, Brown GW. A realist review to assess for whom, under what conditions and how pay for performance programmes work in low-and middle-income countries. Social Science & Medicine. 2021 Feb 1;270:113624.
193. Das A, Gopalan SS, Chandramohan D. Effect of pay for performance to improve quality of maternal and child care in low-and middle-income countries: a systematic review. BMC public health. 2016 Dec;16:1-1.
194. Eijkenaar F. Pay for performance in health care: an international overview of initiatives. Medical Care Research and Review. 2012 Jun;69(3):251-76.
195. Patel S. Structural, institutional and organizational factors associated with successful pay for performance programmes in improving quality of maternal and child health care in low and middle income countries: a systematic literature review. Journal of Global Health. 2018 Dec;8(2).
196. Miller G, Babiarz KS. Pay-for-performance incentives in low-and middle-income country health programs. 2013. National Bureau of Economic Research. Working Paper 18932. DOI: 10.3386/w18932
197. Public Policy and Franchising Reproductive Health: Current Evidence and Future Directions. Geneva: World Health Organization; 2006. https://iris.who.int/bitstream/handle/10665/43735/9789241596021_eng.pdf
198. Bennett S, Dakpallah G, Garner P, Gilson L, Nittayaramphong S, Zurita B, Zwi A. Carrot and stick: state mechanisms to influence private provider behaviour. Health policy and planning. 1994 Mar 1;9(1):1-3.
199. Mahendradhata Y. The case for stronger regulation of private practitioners to control tuberculosis in low-and middle-income countries. BMC Research Notes. 2015 Dec;8:1-5.
200. Masunaga Y, Muela Ribera J, Nguyen TT, Tesfazghi K, Peeters Grietens K. In search of the last malaria cases: ethnographic methods for community and private-sector engagement in malaria elimination in Vietnam, Laos, and Cambodia. Malaria Journal. 2021 Dec;20:1-3.
201. Sidibe A, Maglior A, Cueto C, Chen I, Le Menach A, Chang MA, Eisele TP, Andrinopolous K, Cherubin J, Lemoine JF, Bennett A. Assessing the role of the private sector in surveillance for malaria elimination in Haiti and the Dominican Republic: a qualitative study. Malaria Journal. 2019 Dec;18:1-4.
202. Engaging private health care providers in TB care and prevention: a landscape analysis; Second Edition. Geneva: World Health Organization; 2021. https://iris.who.int/bitstream/handle/10665/351023/9789240027039-eng.pdf?sequence=1
203. Informal Consultation on Malaria and Mobile and Migrant Populations: Addressing Priority Gaps in the Context of Malaria Elimination in the GMS. Bangkok: World Health Organization; 2016. https://iris.who.int/bitstream/handle/10665/258657/RS-2016-GE-71-THA-eng.pdf
204. Axelsson H, Bustreo F, Harding A. Private sector participation in child health: A review of World Bank projects, 1993-2002. World Bank, Washington, DC; 2003.
205. Madhavan S, Bishai D. Private Sector Engagement in Sexual and Reproductive Health and Maternal and Neonatal Health. A Review of the Evidence. Bamenda: Human Development Resource Centre. 2010 Dec 20. https://assets.publishing.service.gov.uk/media/57a08af5ed915d622c0009e9/Private-Sector-Engagement-in-SRH-MNH.pdf
206. Olu O, Drameh-Avognon P, Asamoah-Odei E, Kasolo F, Valdez T, Kabaniha G, Karamagi H, Good S, O’Malley H, Yoti Z, Razakazoa N. Community participation and private sector engagement are fundamental to achieving universal health coverage and health security in Africa: reflections from the second Africa health forum. InBMC proceedings 2019 Nov (Vol. 13, pp. 1-11). BioMed Central.
207. Whyle EB, Olivier J. Models of public–private engagement for health services delivery and financing in Southern Africa: a systematic review. Health policy and planning. 2016 Dec 1;31(10):1515-29.
208. Awor P, Peterson S, Gautham M. Delivering child health interventions through the private sector in low and middle income countries: challenges, opportunities, and potential next steps. bmj. 2018 Jul 30;362.
209. Banu S, Haque F, Ahmed S, Sultana S, Rahman MM, Khatun R, Paul KK, Kabir S, Rahman SM, Banu RS, Islam MS. Social Enterprise Model (SEM) for private sector tuberculosis screening and care in Bangladesh. PLoS One. 2020 Nov 23;15(11):e0241437.
210. Bhatia R, Rastogi RM, Ortega L. Malaria successes and challenges in Asia. Journal of vector borne diseases. 2013 Oct 1;50(4):239-47.
211. Atkinson JA, Vallely A, Fitzgerald L, Whittaker M, Tanner M. The architecture and effect of participation: a systematic review of community participation for communicable disease control and elimination. Implications for malaria elimination. Malaria journal. 2011 Dec;10:1-33.
212. Bloom G, Standing H, Lucas H, Bhuiya A, Oladepo O, Peters DH. Making health markets work better for poor people: the case of informal providers. Health policy and planning. 2011 Jul 1;26(suppl_1):i45-52.
213. Fernando SD, Dharmawardana P, Epasinghe G, Senanayake N, Rodrigo C, Premaratne R, Wickremasinghe R. Contribution of the private sector healthcare service providers to malaria diagnosis in a prevention of re-introduction setting. Malaria Journal. 2016 Dec;15:1-7.
214. Feachem RG, Phillips AA, Hwang J, Cotter C, Wielgosz B, Greenwood BM, Sabot O, Rodriguez MH, Abeyasinghe RR, Ghebreyesus TA, Snow RW. Shrinking the malaria map: progress and prospects. The Lancet. 2010 Nov 6;376(9752):1566-78.
215. Herberholz C, Supakankunti S. Contracting private hospitals: experiences from Southeast and East Asia. Health Policy. 2015 Mar 1;119(3):274-86.
216. James JE. Can public financing of the private sector defeat antimicrobial resistance?. Journal of Public Health. 2019 Jun 1;41(2):422-6.
217. Sekhri N, Feachem R, Ni A. Public-private integrated partnerships demonstrate the potential to improve health care access, quality, and efficiency. Health affairs. 2011 Aug 1;30(8):1498-507.
218. Montagu D, Anglemyer A, Tiwari M, Drasser K, Rutherford GW, Horvath TH, Kennedy GE, Bero L, Shah N, Kinlaw H. Private versus public strategies for health service provision for improving health outcomes in resource limited settings: a systematic review.
219. Corley AG, Thornton CP, Glass NE. The role of nurses and community health workers in confronting neglected tropical diseases in sub-Saharan Africa: a systematic review. PLoS neglected tropical diseases. 2016 Sep 15;10(9):e0004914.
220. Tabrizi JS, Azami-Aghdash S, Gharaee H. Public-private partnership policy in primary health care: a scoping review. Journal of primary care & community health. 2020 Aug;11:2150132720943769.
221. Shrivastava A, Jauhari S, Kumar R. Strengthening of Health Sector through Public Private Partnership (PPP): A Systematic Review. Shodhaytan- AISECT University Journal. 2016. http://aujournals.ipublisher.in/File_upload/89948_92109094.pdf
222. Thadani KB. Public private partnership in the health sector: Boon or bane. Procedia-Social and Behavioral Sciences. 2014 Nov 27;157:307-16.
223. Ullah AN, Huque R, Husain A, Akter S, Islam A, Newell JN. Effectiveness of involving the private medical sector in the National TB Control Programme in Bangladesh: evidence from mixed methods. BMJ open. 2012 Jan 1;2(6):e001534.
224. Roehrich JK, Lewis MA, George G. Are public–private partnerships a healthy option? A systematic literature review. Social science & medicine. 2014 Jul 1;113:110-9.
225. Sajani TT, Alo K, Aktaruzzaman SA. Public Private Partnership (PPP) in health sector of Bangladesh. Anwer Khan Modern Medical College Journal. 2014 May 8;5(1):42-5.
226. Rahman R. Shrinking the state: The rise of private sector healthcare in Bangladesh. Journal of International Development. 2020 Jul;32(5):717-26.
227. Bishai DM, Champion C. The economics of social franchising for health in low-and middle-income countries. InTransforming Health Markets in Asia and Africa 2012 Dec 13 (pp. 162-179). Routledge.
228. Koehlmoos TP, Gazi R, Hossain S, Zaman K. The effect of social franchising on access to and quality of health services in low‐and middle‐income countries. Cochrane database of systematic reviews. 2009(1).
229. Montagu D. Franchising of health services in low-income countries. Health policy and planning. 2002 Jun 1;17(2):121-30.
230. Mills A, Brugha R, Hanson K, McPake B. What can be done about the private health sector in low-income countries?. Bulletin of the World Health Organization. 2002;80:325-30.
231. Hanson K, Gilson L, Goodman C, Mills A, Smith R, Feachem R, Feachem NS, Koehlmoos TP, Kinlaw H. Is private health care the answer to the health problems of the world's poor?. PLoS Medicine. 2008 Nov;5(11):e233.
232. Oxman AD, Fretheim A. Can paying for results help to achieve the Millennium Development Goals? A critical review of selected evaluations of results‐based financing. Journal of Evidence‐Based Medicine. 2009 Aug;2(3):184-95.
233. Mills A, Broomberg J. Experiences of contracting: an overview of the literature. World Health Organization: Geneva. 1998. https://iris.who.int/bitstream/handle/10665/64773/WHO_ICO_MESD_33.pdf
234. Marek T, O’Farrell C, Yamamoto C, Zable I. Trends and opportunities in public-private partnerships to improve health service delivery in Africa. Human Development Sector, Africa Region, The World Bank; 2005 Aug 1. https://documents1.worldbank.org/curated/en/494201468193141531/pdf/531330WP0trent10Box345596B01PUBLIC1.pdf
235. Dixit A, Lee MC, Goettsch B, Afrane Y, Githeko AK, Yan G. Discovering the cost of care: consumer, provider, and retailer surveys shed light on the determinants of malaria health-seeking behaviours. Malaria journal. 2016 Dec;15:1-1.
